# Ventilation and detection of airborne SARS-CoV-2: elucidating high-risk spaces in naturally ventilated healthcare settings

**DOI:** 10.1101/2021.06.30.21258984

**Authors:** Ashley Styczynski, Caitlin Hemlock, Kazi Injamamul Hoque, Renu Verma, Chris LeBoa, Md. Omar Faruk Bhuiyan, Auddithio Nag, Md. Golam Dostogir Harun, Mohammed Badrul Amin, Jason R. Andrews

## Abstract

**Background:** In healthcare settings in low- and middle-income countries, which frequently rely upon natural ventilation, the risk of aerosol transmission of SARS-CoV-2 remains poorly understood. We aimed to evaluate the risk of exposure to SARS-CoV-2 in naturally-ventilated hospital settings by measuring parameters of ventilation and comparing these findings with results of bioaerosol sampling.

**Methods:** We measured outdoor and room CO_2_ to estimate absolute ventilation (liters per second [L/s]) from 9 hospitals in Bangladesh during October 2020 - February 2021. We estimated infectious risk across different spaces using a modified Wells-Riley equation. We collected air samples from these same spaces at 12.5 L/min over 30 minutes and performed RT-qPCR to detect SARS-CoV-2 N-gene. We used multivariable linear regression and calculated elasticity to identify characteristics associated with ventilation.

**Results:** Based on ventilation of 86 patient care areas and COVID-19 case numbers, we found that over a 40-hour exposure period, outpatient departments posed the highest median risk for infection (5.4%), followed by COVID intensive care units (1.8%). We detected SARS-CoV-2 RNA in 18.6% (16/86) of air samples. Ceiling height and total open area of doors and windows were found to have the greatest impact on ventilation.

**Conclusion:** Our findings provide evidence that naturally-ventilated healthcare settings may pose a high risk for exposure to SARS-CoV-2, particularly among non-COVID designated spaces, but improving parameters of ventilation can mitigate this risk.

## BACKGROUND

Hospital-associated exposure to SARS-CoV-2 was a critical driver of disease spread early in the pandemic.[1] Mitigation efforts in healthcare facilities initially focused on reducing droplet and contact exposure to SARS-CoV-2.[2] However, airborne transmission has been increasingly recognized as an important route of virus spread.[3–6] A key component towards reducing the spread of aerosol-transmitted diseases is to ensure adequate ventilation.[7] Many hospitals in high-resource settings isolate patients with COVID-19 in negative pressure rooms with regulated rates of air exchange. These advanced engineering systems are often not available in resource-constrained settings where patients are commonly kept in communal wards that are reliant on natural ventilation. Very little is known about the risk for airborne SARS-CoV-2 transmission in these settings.

The large size of droplet particles limits their spread in both space and time, requiring close proximity to an infected individual to establish exposure. Aerosols, in contrast, can travel in suspended air plumes with prolonged viral persistence.[8,9] Exposure to SARS-CoV-2 through aerosols can thus occur over larger space and time parameters, posing greater cumulative risk in shared spaces with air recirculation and/or inadequate ventilation. Several studies have detected SARS-CoV-2 RNA in air samples from hospitalized patients with COVID-19, including up to 13 feet away from infected patients, demonstrating that the virus is carried in aerosols. ^[10–17]^ Two studies used culture to demonstrate the presence of viable virus in aerosol samples, further supporting airborne transmission as a potential pathway of exposure.[10,18]

With the ongoing transmission of COVID-19 globally, governments have struggled to protect one of the most vulnerable populations: healthcare workers. Large numbers of healthcare worker infections have further burdened healthcare systems, particularly in low- and middle-income countries (LMICs) where dire shortages of healthcare workers preceded the COVID-19 pandemic.[19–21] Adequately protecting healthcare workers relies on understanding transmission risk to inform risk mitigation strategies - from designing engineering controls to implementing appropriate personal protective equipment (PPE) recommendations. Aerosolized particles produced in such environments without adequate ventilation mechanisms could be a critical exposure pathway for healthcare providers. A more complete understanding of the risk of airborne SARS-CoV-2 transmission is imperative for developing policies and practices that improve the safety of healthcare facilities.

World Health Organization guidelines for natural ventilation for infection control in healthcare settings recommends a ventilation rate of 60 liters per second per person (L/s/p) for general wards and outpatient departments (OPD) to prevent airborne infections.[22] Obtaining these ventilation parameters depends on ensuring adequate air exchange efficiency, which is dictated by factors such as opening area to outside, cross-ventilation, and person density.[23,24] A useful metric for approximating ventilation in a steady-state indoor environment is by measuring CO_2_, which reflects the amount of air that is exhaled breath based on the number of people in a given space.[25] This can be translated into infectious risk for airborne infections when the pathogen abundance in the environment can be estimated or modeled.[26,27]

The objective of this study was to quantify the potential risk of SARS-CoV-2 infection across naturally-ventilated healthcare spaces in Dhaka, Bangladesh and to compare these findings with SARS-CoV-2 RNA detection in aerosols in those spaces. We also evaluated the association between ventilation and the presence of SARS-CoV-2 RNA and analyzed drivers of ventilation to identify potential factors to modify the risk in those spaces.

## METHODS

### Ethics

The current study was approved by the research and ethics review committees at icddr,b (#PR-20063). Because the study did not involve a human subjects component, it was considered not to require IRB approval at Stanford University or University of California Berkeley.

### Study Setting

We collected ventilation measurements and conducted environmental bioaerosol sampling in six public and three private hospitals in Dhaka, Bangladesh between October 2020 and February 2021. We selected naturally-ventilated rooms for sampling, which were categorized by whether patients in that area were known or suspected to have COVID-19 or not. We included a range of room types across facilities, including open wards, intensive care units (ICUs), OPDs, PPE doffing areas, and bathrooms.

### Data Collection

We collected measures of ventilation in each of the air sample collection environments to use in our risk modeling. We used a handheld carbon dioxide meter (Extech, Boston, MA) to assess levels of CO_2_ in parts per million (ppm) at 5 minute intervals across the 30-minute sampling period, as well as temperature and humidity. We averaged the CO_2_ levels across the sampling period to approximate a steady-state concentration. We also collected outdoor CO_2_ measurements at the beginning of each day of sample collection. We used these values to calculate the absolute ventilation (L/s) per sampling space using the following equation:[25]

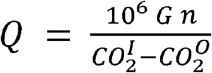

Where *G* is equal to the average CO_2_ generation rate per person, *n* is equal to the number of people in the space, 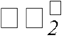 is equal to the averaged CO_2_ measurements during the sampling period, and 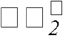 is equal to the outdoor CO_2_ measurement taken in the morning of each sampling day. Ventilation rate (L/s/p) was calculated by dividing the absolute ventilation by the number of people in the space. We also calculated air changes per hour (ACH) using the equation:

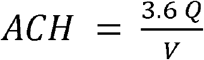

Where *V* is equal to the room volume (m^3^).

We also collected information on the number of COVID (confirmed or suspected) and non-COVID patients, healthcare staff, and visitors present throughout the sample collection time to inform our risk models. For our ventilation analysis, we used a GLM 15 Compact Laser Measure (Bosch, Farmington Hills, MI) to measure room height, width, and length and the area of all open windows and doors. We considered a room to have cross ventilation if there was a window or door open on two opposing walls. Within each of the spaces, we recorded the proportion of patients wearing face masks.

### Risk Modeling

We use the modified Wells-Riley equation proposed by Rudnick and Milton to generate the probability of infection over a 40-hour time interval to represent risk during an average work week.[27]

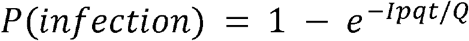

Where *I* is the number of infected individuals in the space, *p* is the average respiration rate of an adult (6 L/min), *t* is the time elapsed in an interval, and *q* is the quantum emission rate (QER), which accounts for the number of infectious doses emitted by an individual over a given time.

For *I*, we counted all patients in designated COVID spaces as potential infectors since a positive COVID test is required for admission to a COVID-designated area. Non-COVID spaces included open wards where all patients have tested negative, and OPDs, where the number of infected individuals is unknown. We set *I* equal to 1 for non-COVID open wards to represent a hypothetical scenario if one patient were to be incorrectly diagnosed as negative. For OPDs, we assumed 3% of non-staff were infected based on concurrent SARS-CoV-2 test positivity data for the given sampling period (rounding up for fractional values).[28]

To obtain *q*, we used activity-specific distributions characterized by Buonanno et al.[29] The authors hypothesized that there is uncertainty around the QER due to random variation in the concentration of viral load expired during activity. For ICUs, open wards, and private rooms, we assumed the activity level was “resting, oral breathing” for potential infectors [log_10_(QER per hour) ∼ N(−0.429, 0.720)] and for OPDs, we assumed “light activity, talking” for potential infectors [log_10_(QER per hour) ∼ N(0.698, 0.720)]. We applied a Monte Carlo method to draw values for each potential infector QER within each space (*q*_*i*_). Based on studies reporting time to diagnosis and hospitalization for SARS-CoV-2, we estimated that inpatients were on average 8 days into their disease course compared with 3 days for outpatients.[30–32] Since viral shedding decreases with duration of illness, we thus subtracted 0.5 (log_10_ scale) from each *q*_*i*_ drawn for inpatients based on estimated differences in viral shedding between day 3 and day 8 of illness.[33,34] We calculated the average *q*_*i*_ drawn for all potential infectors, took the antilog, and multiplied by one minus the proportion of mask-wearing patients (*P*_*m*_) times the efficacy of surgical masks (*E*) in preventing outward transmission of infectious aerosols (∼70%) to obtain the final value of *q* for each sampling space.[35,36]

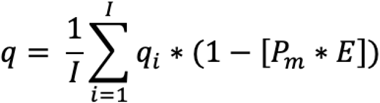

Using the final calculated value of *q* for each space, we calculated the risk for each sampling space using the aforementioned modified Wells-Riley equation and took the median risk for each type of space (e.g. to obtain the median risk for OPDs). We repeated random draws of *q*_*i*_ for potential infectors 1,000 times to obtain a distribution of 1,000 type-specific medians. We calculated the overall median risk by type of space for the 1,000 simulations, as well as the 2.5th percentile and 97.5th percentile of the generated distribution. We also used the 1000 calculated *q* values to obtain risk curves for individual sampling spaces.

### Bioaerosol Sampling

To compare the results of our risk modeling to empirical data, we collected air samples over a 30-minute time period in each space and measured SARS-CoV-2 viral copies.

#### Sample Collection

We used an SKC biosampler liquid impinger with a BioLite Air sampling pump (SKC Inc., Eighty Four, PA) set to a calibrated flow rate of 12.5 L/minute. The biosampler was set as close to the center of each room as possible and 1-1.5 m above the ground. The vessel connected to the liquid impinger was filled with 10 mL of 1x PBS (pH 7.4) for bioaerosol collection. Immediately after the collection, we added 7 mL of NucliSENS® RNA stabilizing lysis buffer (bioMérieux, Inc., Durham, NC) and transported the samples at 2-8°C. Between each sample collection, we decontaminated the biosampler by separating all components of the device and submerging them into 10% sodium hypochlorite for 15 minutes followed by rinsing the sampler components thrice with distilled water. To rule out any carryover contamination from the previous run, we rinsed the biosampler after every decontamination with 1 mL distilled water that we analyzed by RT-qPCR. Additionally, to ensure that there was no backflow contamination by the pump, we collected negative controls daily by attaching an N95 filter to the biosampler inlet over a 30-minute sampling period and tested the PBS collection fluid by RT-qPCR.

#### Sample Processing and RNA Extraction

Before RNA extraction, we concentrated 14 mL of sample collected in PBS and lysis buffer to 500 μL using Amicon® Ultra-15 Centrifugal Filter Units (Milipore Sigma, cat# C7715) at 5000 rpm for 20 minutes. We extracted RNA using a modified (additional 25 μL of proteinase K was added to the reaction during the lysis step) MagMAX Viral/Pathogen Ultra Nucleic Acid Isolation Kit (Applied Biosystems A42356) as per the manufacturer’s instructions. The RNA was eluted in a 50 μL elution buffer and stored at -20°C until further testing.

#### Sample Analysis

We performed RT-qPCR using the CDC qualified primers (500 nM) and TaqMan probes (300 nM) amplifying N1 and N2 regions of SARS-CoV-2 N-gene.[37] TaqPath one-step RT-qPCR mastermix (Invitrogen, Darmstadt, Germany) was used in a 20 μL reaction volume and analyzed on a StepOne-Plus (Applied Biosystems) instrument, using the following program: 10 min at 50°C for reverse transcription, followed by 3 min at 95°C and 40 cycles of 10 s at 95°C, 15 s at 56°C, and 5 s at 72°C. We estimated the number of copies per sample from a standard curve using 10-fold serially diluted SARS-CoV-2 synthetic RNA (ATCC Cat#VR-3276SD). All the samples were run in duplicates and averaged. A positive sample was defined as having a Ct value less than 38 in at least two measurements (N1 target positive in both replicates, N2 target positive in both replicates, or positive N1 target + positive N2 target).

### Ventilation Analysis

To determine which parameters of ventilation had the greatest impact on ventilation, we analyzed the association between each ventilation parameter measured and log_10_ absolute ventilation using univariate linear regression models to obtain unadjusted mean differences, excluding extreme outliers from each parameter distribution. To alleviate sample size constraints, we used LASSO regression to select parameters for a multivariable model. We used 5-fold cross-validation to select the λ penalty at the minimum mean-squared error. Any variables with non-zero coefficient values were included in a multivariable linear regression model to obtain adjusted mean differences and 95% confidence intervals.

We assessed variable importance using elasticity, which standardizes the estimated parameters by multiplying the coefficient from the regression model with the mean of the associated variable divided by the mean of the outcome. This results in an estimate of the percent change in the outcome for a percent change in the exposure and makes the parameters comparable in a post-estimation step, while keeping the regression coefficients in units that are understandable.[38]

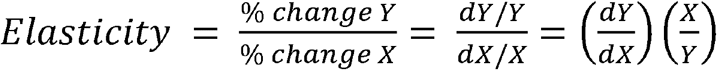

Where *dY/dX* is the slope of *Y* with respect to *X* for a given *X*, multiplied by the ratio of a given value of *X* to a given value of *Y* (we used the mean of both). We performed this for the point estimates and associated confidence intervals from our multivariable regression to identify variables that most influenced log_10_ absolute ventilation. To assess risk stratified by important variables, we employed the approach detailed above and performed 1,000 simulations on each space to generate sample-specific risk medians.

## RESULTS

We sampled a total of 86 locations, including 28 open wards, 9 ICUs, 18 private rooms, 12 OPDs, and 19 other spaces, including bathrooms, PPE doffing rooms, COVID-19 testing areas, and a canteen. Among the 86 spaces, 65 (76%) were areas with patients confirmed or suspected to have COVID-19, and 21 (24%) were non-COVID areas. The areas with COVID-19 patients had fewer people (median: 6) compared with non-COVID areas (median: 38) (p-value <0.001). Within OPDs, the median percent of people found to be wearing masks was 75% (range: 31 - 100%) with the vast majority wearing surgical masks. No hospitalized patients were observed to be wearing masks. Healthcare staff in COVID ICUs wore N95s while staff in other settings typically wore either surgical masks or no masks.

OPDs had the highest person density of the spaces sampled (0.33 people per m^2^), followed by open wards (0.19 people per m^2^) (Table 1). Most rooms apart from wards did not have open windows; the mean open window area of rooms with open windows was 0.49 m^2^. However, the majority of sampled spaces (79%) had at least one open door at the time of sampling. Less than a third (30%) of the spaces had evidence of cross-ventilation. Approximately one-third of spaces used fans for climate control, while ICUs and private rooms predominantly relied on wall or portable air conditioning units.

**Table 1.** Descriptive parameters of sampled spaces (n=86)

|  | ICU<br>(n=9) | OPD<br>(n=12) | Open ward<br>(n=28) | Other*<br>(n=19) | Private room<br>(n=18) | Overall<br>(n=86) |
| --- | --- | --- | --- | --- | --- | --- |
| Room volume (m<sup>3</sup>) | 332<br>[263, 523] | 251<br>[149, 428] | 278<br>[129, 1,180] | 29.3<br>[16.5, 73.6] | 82.1<br>[61.3, 91.7] | 133<br>[70, 344] |
| Floor area (m<sup>2</sup>) | 111.4<br>[78.9, 172.8] | 94.16<br>[54.4, 159] | 82.63<br>[38.8, 390] | 12.01<br>[6.6, 21.6] | 32.34<br>[23, 33.7] | 48.45<br>[22.6, 111] |
| Ceiling height (m) | 2.60<br>[2.46, 3.45] | 2.72<br>[2.69, 3.15] | 3.23<br>[2.67, 3.43] | 2.67<br>[2.41, 3.38] | 2.57<br>[2.54, 3.01] | 2.76<br>[2.55, 3.40] |
| Number of People<br>in Room | 11.33<br>[10.7, 18.3] | 45.7<br>[16.3, 74.8] | 24.0<br>[9.00, 47.3] | 1.33<br>[1.00, 2.42] | 2.50<br>[2.00, 3.25] | 8.00<br>[2.70, 25.1] |
| Person density (per<br>floor area, m<sup>2</sup>) | 0.11<br>[0.07, 0.16] | 0.33<br>[0.24, 0.66] | 0.19<br>[0.08, 0.28] | 0.08<br>[0.01, 0.11] | 0.08<br>[0.07, 0.12] | 0.13<br>[0.07, 0.24] |
| Open window area<br>(m<sup>2</sup>) | 0.00<br>[0.0, 0.0] | 0.00<br>[0.0, 1.24] | 0.49<br>[0.00, 3.00] | 0.0<br>[0.0, 0.0] | 0.0<br>[0.0, 0.0] | 0.0<br>[0.0, 0.8] |
| Open door area (m<sup>2</sup>) | 0.00<br>[0.00, 2.16] | 4.2<br>[3.36, 14.5] | 3.10<br>[1.88, 5.16] | 1.55<br>[0.00, 1.76] | 1.64<br>[0.21, 2.35] | 2.14<br>[1.29, 3.37] |
| Total Open Area<br>(m<sup>2</sup>) | 0.00<br>[0.00, 2.16] | 6.61<br>[4.15, 17.2] | 4.19<br>[2.99, 8.36] | 1.55<br>[0.00, 2.37] | 2.12<br>[1.14, 2.35] | 0.00<br>[0.00, 2.16] |
| Open area to<br>Volume Ratio | 0.00<br>[0.00, 0.01] | 0.04<br>[0.01, 0.07] | 0.01<br>[0.01, 0.02] | 0.01<br>[0.00, 0.04] | 0.02<br>[0.01, 0.03] | 0.00<br>[0.00, 0.01] |
| Any fans in<br>operation | 3<br>(33.3) | 3<br>(25.0) | 14<br>(50.0) | 1<br>(5.3) | 6<br>(33.3) | 27<br>(31.4) |
| Any A/C in<br>operation | 7<br>(77.8) | 1<br>(8.3) | 2<br>(7.1) | 4<br>(21.1) | 7<br>(38.9) | 21<br>(24.4) |
| Cross ventilation<br>present | 2<br>(22.2) | 3<br>(25.0) | 16<br>(57.1) | 4<br>(21.1) | 1<br>(5.6) | 26<br>(30.2) |
| Steady state CO2<br>(ppm) | 931<br>[793, 1,145] | 1,221<br>[835, 1,493] | 765<br>[511, 860] | 892<br>[666, 1,272] | 617<br>[536, 763] | 809<br>[576, 1,126] |
| Absolute<br>ventilation (L/s) | 171<br>[87, 180] | 354<br>[105, 423] | 293<br>[193, 831] | 32<br>[6.83, 43.6] | 50.6<br>[37.1, 137] | 163<br>[49.9, 363] |
| Ventilation per<br>person (L/s/p) | 9.79<br>[7.03, 13.38] | 6.63<br>[4.80, 12.3] | 13.9<br>[11.4, 42.4] | 11.4<br>[6.71, 22.9] | 24.9<br>[14.7, 37.0] | 13.0<br>[7.91, 27.8] |
| ACH | 1.66<br>[1.21, 1.98] | 4.54<br>[2.05, 7.40] | 3.64<br>[2.73, 5.13] | 2.17<br>[1.09, 4.93] | 2.94<br>[1.53, 5.50] | 2.94<br>[1.63, 5.44] |

\*Other spaces include bathrooms (n=11), COVID-19 testing areas (n=4), PPE doffing rooms (n=3), and a canteen (n=1). Data with brackets indicates the median and [IQR] of the variable. Data with parentheses indicates the count and (%) of rooms where the variable is present.

Among the sampled spaces, the median CO_2_ level was 809 ppm (range: 403 ppm - 3166 ppm). CO_2_ values were fairly steady over the 30 minute sampling period (Supplementary Figure 1). The median absolute ventilation among spaces where ventilation was able to be calculated (excluding 6 spaces with no people present during the sampling period) was 163 L/s (IQR 49.9-363) and the ventilation rate was 13.0 L/s/p (IQR 7.9-27.8) (Table 1). An overwhelming majority (83.8%) of patient care rooms had ventilation rates that fell short of the recommended ventilation rate of 60 L/s/p (Figure 1).

**Figure 1.**
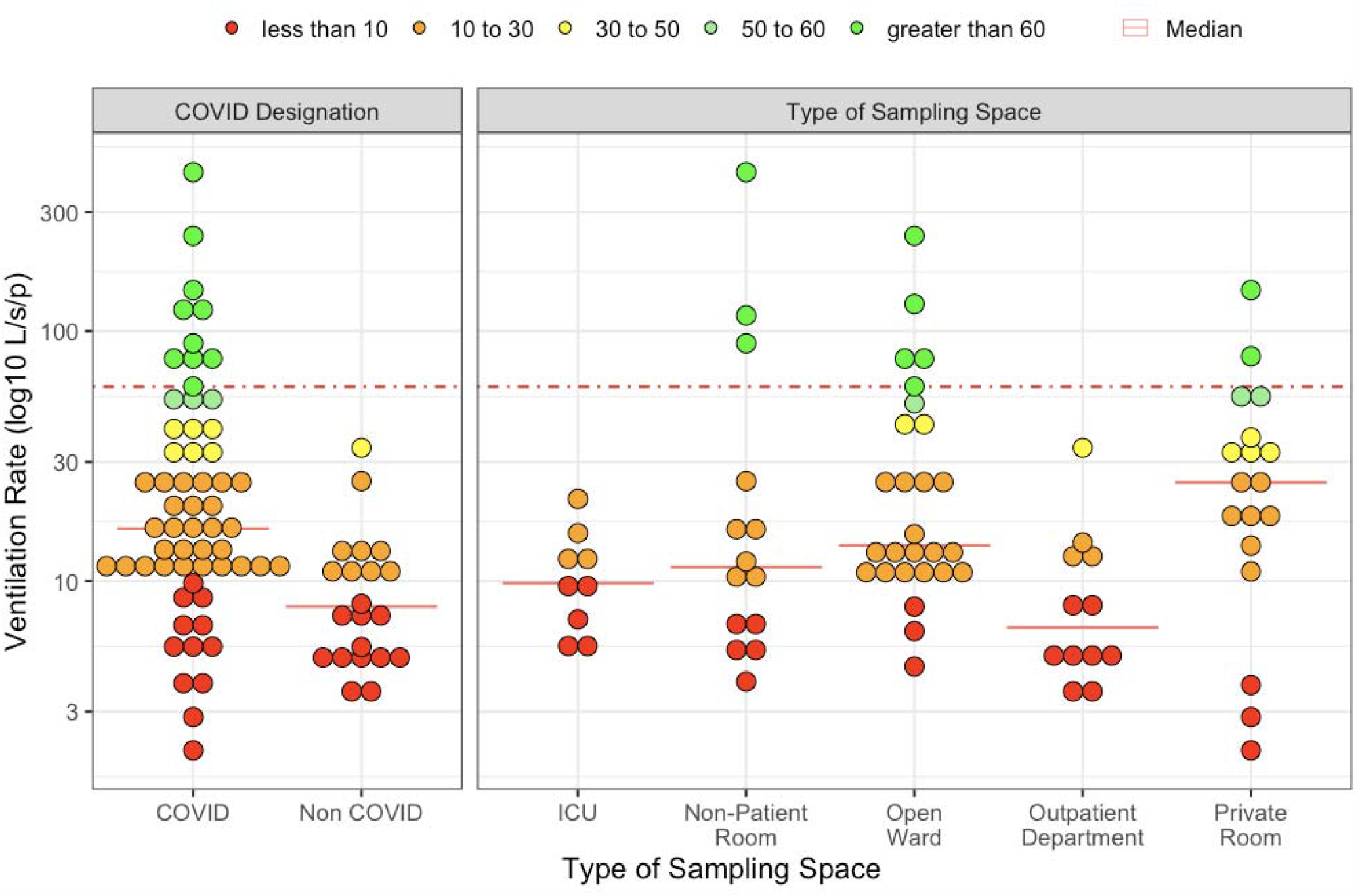
Ventilation rates across sampled patient care areas (n=80), excluding 6 spaces where no people were present at the time of sampling. Dashed line indicates the recommended ventilation rate of 60 L/s/person. Green dots (n=13) signify sampled locations above the 60 L/s/person threshold; yellow dots represent ventilation rates of 30-50 L/s/person (n=7); orange dots represent 10-30 L/s/person (n=36); and red dots are below 10 L/s/person (n=24).

### Estimating Infectious Risk

On average, COVID ICUs and COVID open wards had the highest number of potential infectors (median = 6 for both), compared with COVID private rooms, non-COVID open wards, and OPDs (median = 1 for all). However, we found that among the five types of patient care spaces sampled, OPDs were overall the highest risk location (Figure 2). After 40 hours in OPDs, the median risk of infection in the absence of other mitigation measures was 5.4% (95% CI: 2.1%, 13.1%). ICUs designated as COVID spaces were the second riskiest spaces, with 1.8% risk (95% CI: 0.9%, 3.2%) over 40 hours. Private rooms and open wards for patients with COVID-19 had a similar risk profile (1.1%; 95% CI: 0.5%, 2.7% compared with 1.1%; 95% CI: 0.8%, 3.2%). Open wards that were not designated for patients with COVID-19 carried the least risk under the assumed scenario of one potential infector (0.1%; 0.0% - 0.6%).

**Figure 2.**
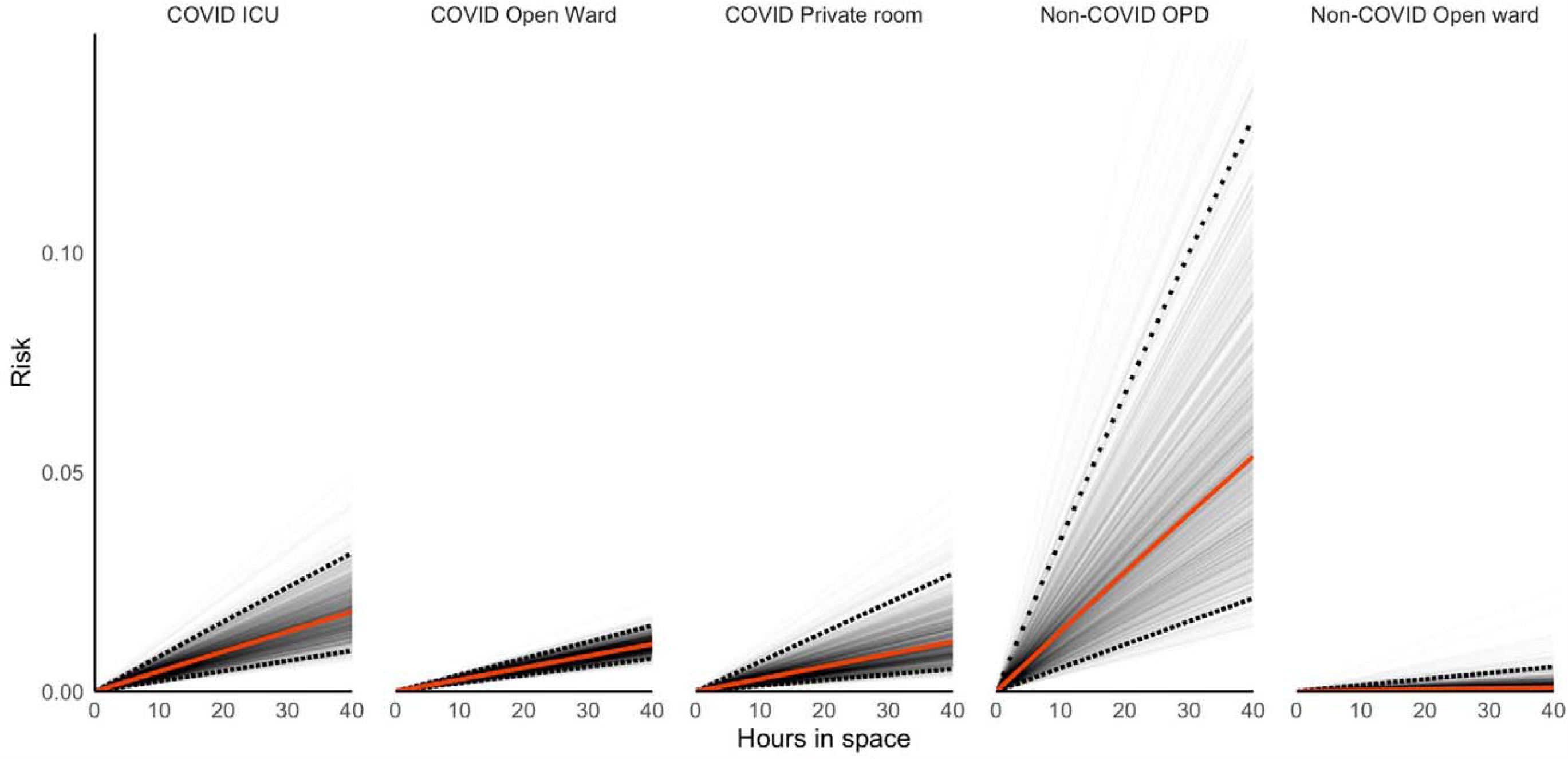
Risk of SARS-CoV-2 infection over a 40-hour time period, by type of sampling space. Gray lines are simulation-specific averages of the median risk by type of sampling space. The solid red line is the overall median risk of infection over time and the dashed black lines are the 2.5th and 97.5th percentiles.

### SARS-CoV-2 RNA Detection

Of the 86 bioaerosol samples tested by RT-qPCR, we detected SARS-CoV-2 RNA in 16 (18.6%) of the samples (Table 2). The room types with the highest proportion of positive samples were non-COVID wards (2/4; 50%) and OPDs (3/12; 25%). Among positive samples, the median copy number was 189 (range: 79-929). There was no difference in the median copy number between COVID and non-COVID spaces where SARS-CoV-2 was detected (p=0.336).

**Table 2.**
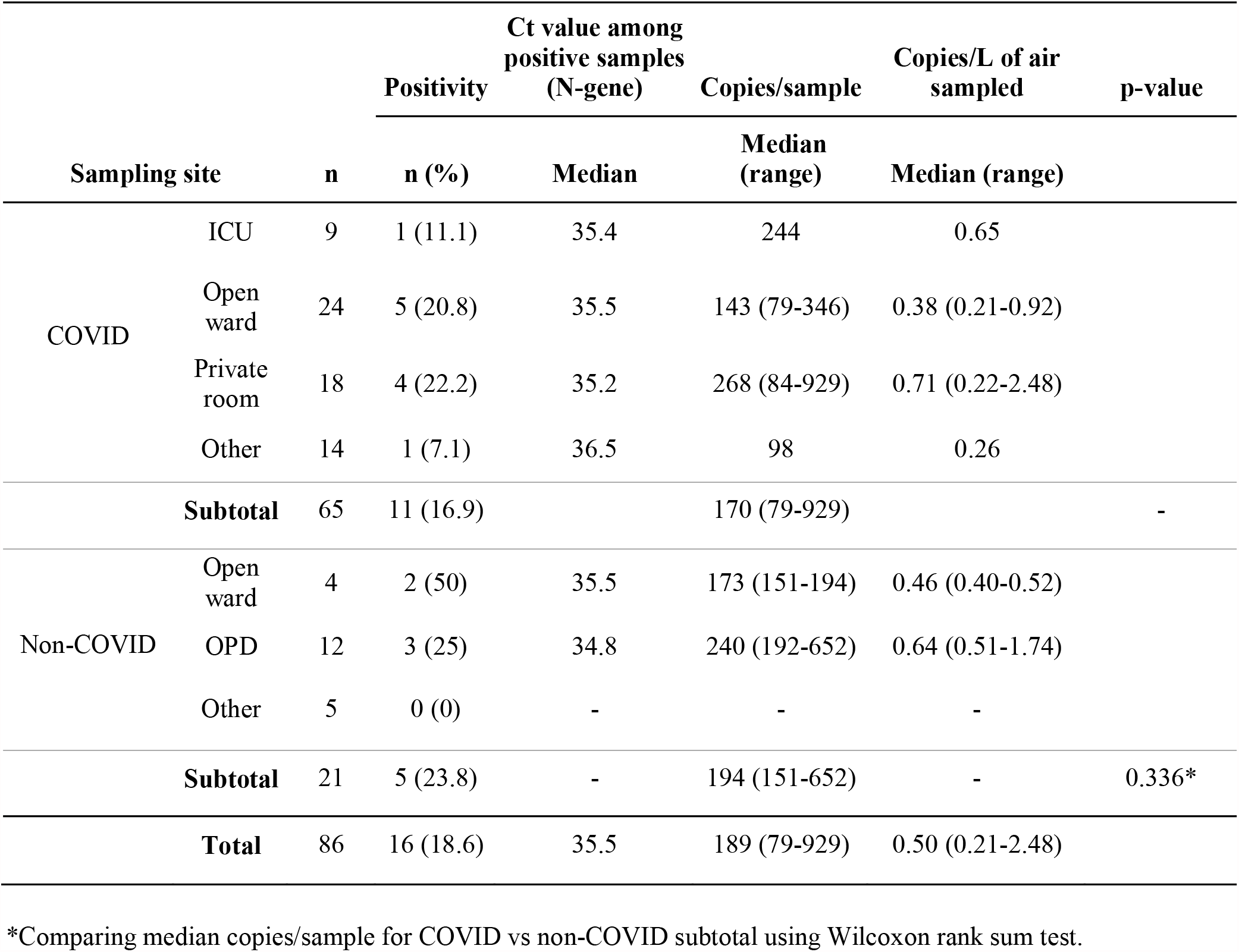
SARS-CoV-2 RNA RT-qPCR positivity and copy numbers in bioaerosol samples collected from nine hospitals in Dhaka, Bangladesh.

### Ventilation Analysis

When analyzing the architectural and ventilation features of the sampling spaces, we found that ceiling height was the most important parameter associated with absolute ventilation in a multivariable analysis with a 6.3% change in log_10_ ventilation per 10% change in ceiling height (95% CI: 3.2%, 9.4%) (Table 3, Supplementary Figure 2). Total open area of doors and windows combined and floor area were the next most influential factors, however these estimates were only marginally significant (0.7%; 95% CI: 0.0%, 1.4% and 0.4%; 95% CI: -0.1%, 0.8%). Having an air conditioning unit running was the next most important parameter (0.3%; 95% CI: 0.0%, 0.6%). Cross ventilation was less important than these factors and was not significant in the multivariable analysis (0.1%; 95% CI: -0.2%, 0.4%). We found the risk of SARS-CoV-2 infection was moderated by total open areas of doors and windows and ceiling height (Supplementary Figure 3). When stratified by ceiling height, doubling the total open area of doors and windows in OPDs reduces the risk of SARS-CoV-2 infection by nearly 50%.

**Table 3.** Univariable, multivariable, and elasticity analysis of ventilation parameters

|  | Unadjusted mean difference<br>(95% CI) | LASSO<br>coefficient<sup>1</sup> | Adjusted mean difference<br>(95% CI)<sup>1</sup> | Elasticity<sup>2</sup> |
| --- | --- | --- | --- | --- |
| Average number of people | 0.014 (0.01, 0.018) | 0.0037 | 0.003 (-0.00088, 0.0069) | 0.3% (-0.089%, 0.7%) |
| **Architectural features** |  |  |  |  |
| Ceiling height (m) | 0.43 (0.098, 0.77) | 0.4 | 0.44 (0.22, 0.66) | 6.3% (3.2%, 9.4%) |
| Room volume (m<sup>3</sup>) | 0.00096 (7e-04, 0.0012) | 0 | - | - |
| Floor area (m<sup>2</sup>) | 0.0025 (0.0018, 0.0032) | 0.00055 | 0.00065 (-0.00016, 0.0015) | 0.35% (-0.086%, 0.79%) |
| **Ventilation features** |  |  |  |  |
| Open door area (m<sup>2</sup>) | 0.095 (0.06, 0.13) | 0.0085 | 0.0085 (-0.039, 0.055) | 0.13% (-0.57%, 0.83%) |
| Open window area (m<sup>2</sup>) | 0.074 (0.046, 0.1) | 0 | - | - |
| Total open window and door area<br>(m<sup>2</sup>) | 0.057 (0.041, 0.073) | 0.037 | 0.036 (-0.00023, 0.072) | 0.7% (-0.0046%, 1.4%) |
| Ratio of open window and door area<br>to floor area | 0.023 (-2.3, 2.4) | 0 | - | - |
| Any AC on | -0.27 (-0.62, 0.092) | 0.17 | 0.24 (0.025, 0.46) | 0.31% (0.032%, 0.59%) |
| Any fans on | 0.76 (0.47, 1.1) | 0.04 | 0.081 (-0.14, 0.31) | 0.12% (-0.22%, 0.47%) |
| Cross ventilation present | 0.8 (0.51, 1.1) | 0.11 | 0.1 (-0.12, 0.32) | 0.13% (-0.16%, 0.43%) |
| **Type of sampling space** |  |  |  |  |
| Private room | ref. | ref. | ref. |  |
| Open ward | 0.94 (0.61, 1.3) | 0.31 | 0.38 (0.13, 0.64) | - |
| ICU | 0.39 (-0.049, 0.82) | 0.15 | 0.18 (-0.13, 0.49) | - |
| OPD | 0.66 (0.27, 1.1) | 0.15 | 0.27 (-0.059, 0.59) | - |
| Other | -0.31 (-0.69, 0.069) | -0.22 | -0.19 (-0.43, 0.05) | - |

<sup>1</sup> Adjusted for temperature, humidity, and whether a window or door was open nearby as nuisance parameters
<sup>2</sup> Interpreted as the percentage change in log<sub>10</sub> absolute ventilation for a 10% increase in the given variable. Note: not estimated for type of sampling space or parameters with LASSO coefficient of 0.

## DISCUSSION

In this study, we demonstrated the potential for airborne transmission of SARS-CoV-2 by revealing inadequate ventilation, a known driver of infectious risk, in a variety of naturally-ventilated healthcare spaces. There was severely low ventilation across most spaces, indicating a need for large-scale improvements to reduce environmental exposure risk.[22]

We found that patient entry points to the hospitals (OPDs) where patient COVID-19 status was unknown had the highest risk for airborne SARS-CoV-2 exposure based on infectious risk modeling, despite high rates of mask wearing. This was corroborated by higher rates of viral RNA detection in these spaces. We believe this is explained by several factors. Patients presenting to OPDs likely have higher viral shedding because they are earlier in their disease course, and they may have a higher level of activity than patients admitted to the hospital, both of which we accounted for in our modeling. This corresponds with findings that the majority of SARS-CoV-2 infections are likely transmitted by people early in their disease course.[39] Additionally, although OPDs had higher absolute ventilation than some other areas, it was still insufficient to mitigate the risk of higher viral shedding. Conversely, despite high numbers of infectious individuals in COVID wards, adequate ventilation, mediated through higher ceilings and open windows/doors, kept the overall risk low. Diverging from our risk models, there was also a high rate of detection in non-COVID wards, potentially due to imperfect testing methods during triage. The high proportion of positive samples in non-COVID spaces is an important finding because healthcare workers may underestimate exposure risk in these settings. This was supported by the lack of N95 masks worn by healthcare workers in these spaces.

Areas with patients confirmed or suspected to have COVID-19 are often considered to be high-risk areas for transmission potential, leading to differential PPE recommendations across types of patient care areas.[40] Additionally, a lower perceived risk of exposure to SARS-CoV-2 has been associated with decreased adherence to PPE use.[41,42] In accordance with this observation, PPE use among healthcare staff in our study appeared to be less stringent in non-COVID areas. Our study revealed a strong dichotomy between perceived risk and actual risk of exposure to SARS-CoV-2 across healthcare spaces, which appears to be heavily modulated by viral emission rate and timing of disease course. These findings demonstrate an urgent need for enhanced ventilation measures across all healthcare settings for the protection of healthcare workers, patients, and visitors against nosocomial transmission of airborne diseases, including COVID-19.

We demonstrated direct detection of SARS-CoV-2 RNA in a high percentage of healthcare spaces - we found SARS-CoV-2 RNA in 18.6% of samples. In contrast, four other studies failed to detect SARS-CoV-2 RNA in mechanically-ventilated healthcare spaces despite obtaining substantially larger volumes of filtered air and placing air samplers proximal to infected patients.[43–46] We also did not detect SARS-CoV-2 in bathrooms or PPE doffing rooms, contrasting with other studies that found these to be high-risk areas, indicating that risk in these spaces may be context dependent.[47]

Of the samples in which we detected SARS-CoV-2 RNA, the median copy number was less than 1 copy/L of air sampled, which is comparable to other studies where SARS-CoV-2 RNA was detected in air samples from healthcare settings.[10–14,48] Given that previous estimates have suggested an infectious dose of SARS-CoV-2 may be in the range of 100-300 virions,[49,50] this could indicate the risk from aerosol transmission may be substantial, especially given that cumulative exposure may be more important for establishing infection than exposure at a single time point.[51] However, it is difficult to translate results of air sample collection to exposure to infectious quanta given limitations of the sample collection process.

A unique aspect of this study is an investigation of which parameters of ventilation have the greatest impact on air exchange in naturally-ventilated settings and subsequently how risk was modified by these factors. Ceiling height was found to be the most influential parameter in the elasticity analysis, which has been previously demonstrated to be an important consideration.[24] However, as this is often not a modifiable factor, it is important to consider the other parameters, such as total open window and door area, which can be particularly influential in OPDs that had the lowest per-person ventilation. Still, considering ceiling height of a room may be important when selecting spaces for OPDs or for choosing spaces to isolate patients with pathogens transmitted via aerosols. Cross ventilation was not noted to be a significant factor affecting ventilation, but this may be because the open area of the windows or doors on opposite walls was not sufficient to overcome the large room volume in the spaces that we sampled.

Our modeled estimates of risk are likely conservative because we did not take into account parameters around aerosol-generating procedures, including the use of positive pressure ventilation, which can further increase risk for airborne transmission. In contrast with other studies,[12,17,52] the ICUs in our study did not have a high rate of SARS-CoV-2 detection. However, inadequate ventilation in these spaces put them as the second highest-risk space in our risk models. Additionally, we assumed maximal effectiveness of masks at preventing outward disease spread by patients based on laboratory studies, though mask use practices (e.g. correct mask wearing over nose and mouth) and mask filtration efficiency are likely to be lower in real-world scenarios compared with standardized study conditions. Furthermore, we identified potential infectors in OPD spaces based on an average test positivity rate at the lowest point in the pandemic in Bangladesh,[28] which coincided with our air sample collection period. However, the risk of infection with SARS-CoV-2 is likely to increase in these spaces as community transmission increases because a higher proportion of patients seeking care are likely to have COVID-19. Therefore, our estimates are conservative for these settings and actual risk during a given time period may be substantially higher.

One limitation of this study is that viral culture was not available. While assessing for the presence of SARS-CoV-2 using RNA does not address pathogen viability, epidemiologic and laboratory studies have demonstrated the possibility of aerosol transmission in the setting of disease spread without direct contact between individuals.[53–56] Additionally, laboratory studies have revealed that the SARS-CoV-2 can remain viable in the air for up to 16 hours with no detectable half life.[9] Furthermore, viability has never been demonstrated in well-known airborne-transmitted diseases such as measles given limitations in collection methods.[57]

Another limitation is the cross-sectional nature of this study, allowing us to only measure SARS-CoV-2 RNA at a single time point, which can be affected by disease constellations, density, and activity of patients. To address this limitation, we combined these direct measures of viral presence with indirect drivers of risk, such as ventilation, to obtain a more complete assessment as ventilation parameters are likely to remain stable over time. This approach to using ventilation parameters as assessed by CO_2_ levels has been validated as a proxy for risk of transmission of other airborne diseases, including tuberculosis and measles.[23,26,27,58]

Given increasing evidence for aerosol transmission of SARS-CoV-2, engineering modifications of healthcare spaces are of utmost importance for reducing risk to healthcare workers, visitors, and other patients of healthcare facilities.[59] Modifications of healthcare spaces were proposed during the first SARS outbreak, including window exhaust fans, that require minimal infrastructure investment and may be relevant in contexts such as Bangladesh and other LMICs.[22,60] Enhanced effects of cross-ventilation may be observed if the open area of windows or doors on opposing walls can be maximized. Additionally, triaging patients in tents outside hospital facilities could result in less virus emission indoors. Barring modifications to the environmental context, reducing the number of people in a space and increased enforcement of surgical mask wearing for patients, especially in OPDs, and N95 use among staff in all patient care areas in the setting of community transmission of COVID-19 may be downstream solutions.[59,61]

## CONCLUSION

COVID-19 remains an ongoing threat to populations around the world. As with outbreaks of other emerging infectious diseases,[62–64] healthcare facilities were a source of transmission in the early spread of SARS-CoV-2, resulting in an excess of healthcare worker infections.[1,20] Improving the safety of healthcare facilities is imperative for protecting against future epidemic spread. This is only possible by adequately equipping healthcare spaces with durable mitigation measures that are effective against a range of transmission patterns. While the COVID-19 pandemic will eventually subside, the risk of airborne transmission of other diseases remains a substantial risk in healthcare facilities with inadequate ventilation. Now is the critical moment of action to prevent healthcare facilities from further amplifying the current and future pandemics.

## Data Availability

All data are available within the manuscript.

## Acknowledgements

We would like to thank Aninda Rahman, Communicable Disease Control Program, Directorate General of Health Services (DGHS), for his facilitation to conduct this research. We would also like to thank Pedro M. de Oliveira, Savvas Gkantonas, and Epaminondas Mastorakos from the University of Cambridge Department of Engineering for sharing thoughtful insights around airborne transmission risk modeling. Additionally, we appreciate the cooperation of the hospitals in Dhaka who allowed us to collect these measurements.

## Competing interests

None.

## Funding

This study was supported by an anonymous donation to the Stanford University School of Medicine. icddr,b acknowledges with gratitude the funding support of Stanford University School of Medicine. icddr,b is also thankful to the following donors: the governments of Bangladesh, Canada, Sweden, and United Kingdom for providing core/ unrestricted support. The donors had no role in data analysis, interpretation, or decision to publish the findings.

## Contributorship

AS contributed to the research conception and design and manuscript writing; CH performed data analysis and contributed to research design and manuscript writing; RV participated in the research design, data analysis and interpretation, and manuscript writing; KIH contributed to the research design, data collection, and data analysis; CL participated in the research design, data analysis, and manuscript writing; AN contributed to research design and data collection; MOFB participated in data collection; MGDH contributed to research design and dissemination of findings for public health practice; MBA contributed to the research conception and design and interpretation of results; JA contributed to research conception and design, interpretation of findings, and manuscript writing.

## SUPPLEMENTARY MATERIALS

**Supplementary Figure 1.**
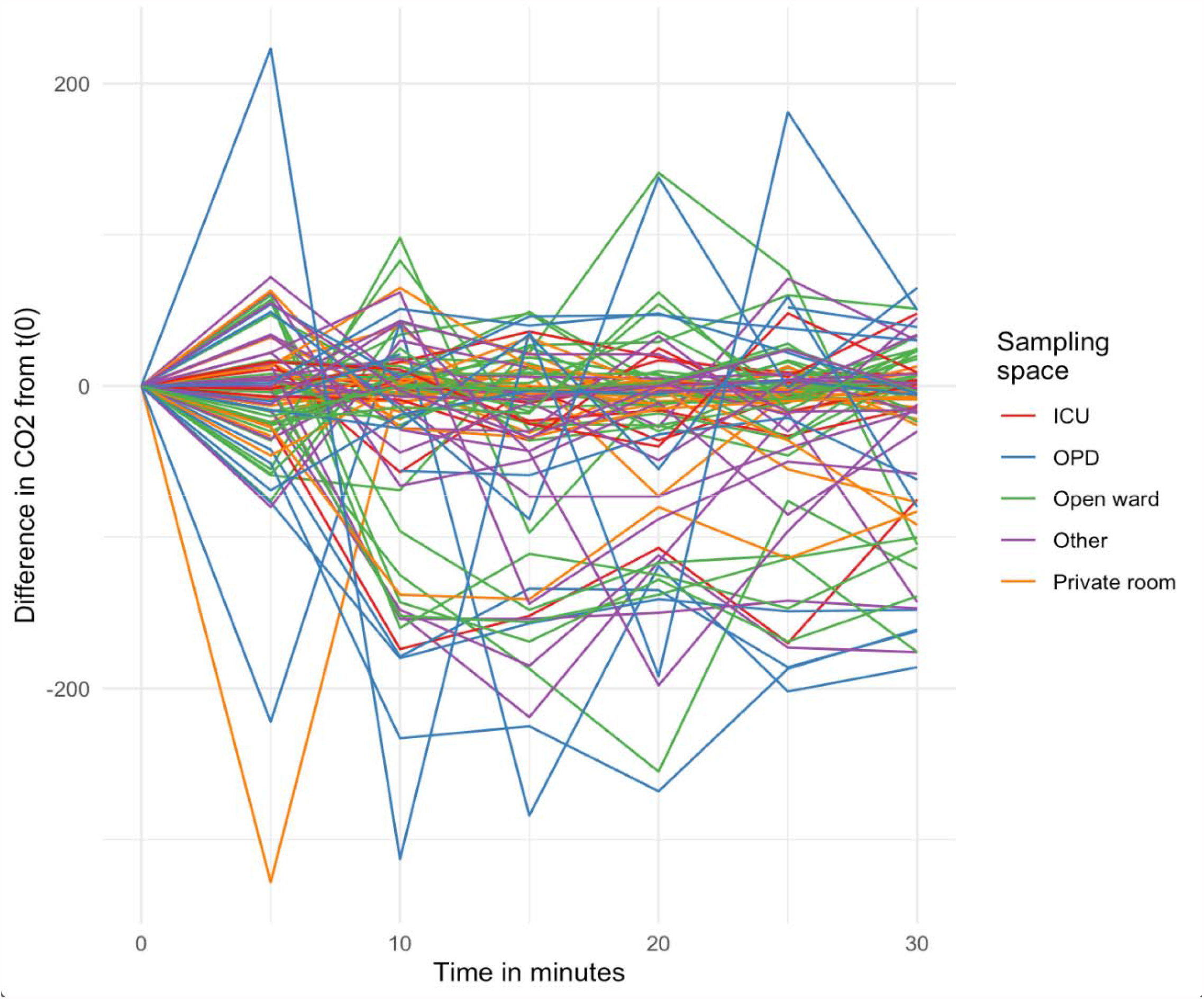
Difference in CO_2_ parts per million (ppm) over the sampling period compared to start time, by type of sampling space Difference in CO_2_ parts per million (ppm) measurements from a given type of sampling space across 5-minute time intervals. ICUs are denoted with red lines; OPDs are represented by blue lines; green lines are open wards; orange lines are private rooms; and purple lines represent other spaces.

**Supplementary Figure 2.**
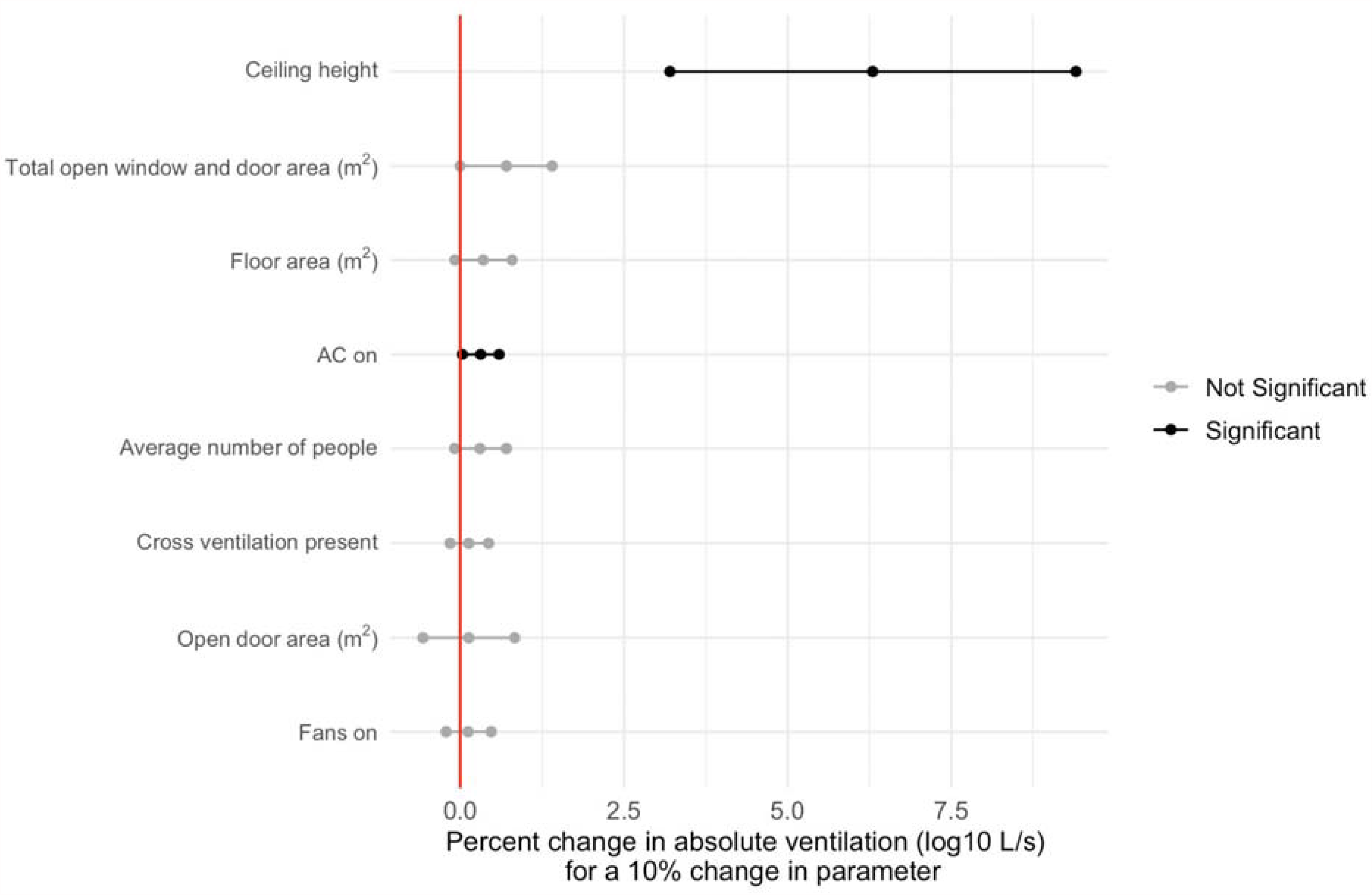
Elasticity of architectural and ventilation features on absolute ventilation (log_10_ L/s) of sampling space (95% confidence interval included) Impact of a 10% change of a given parameter on absolute ventilation. Black lines represent variables that reach statistical significance. Gray lines represent parameters that were not significant.

**Supplementary Figure 3.**
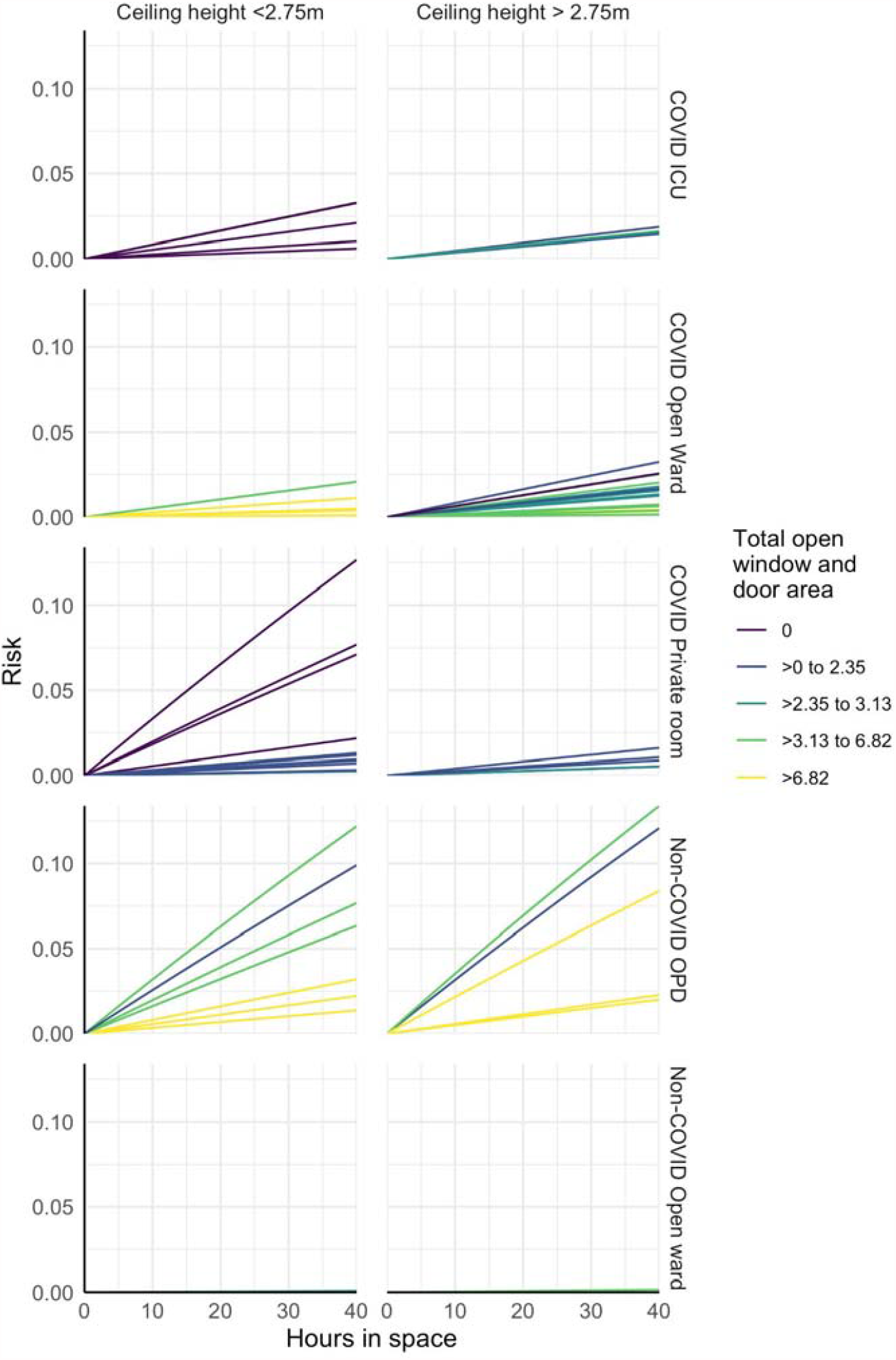
Risk of SARS-CoV-2 infection over 40-hour time period by type of sampling space and ceiling height Cumulative risk of SARS-CoV-2 infection over 40 hours, stratified by ceiling height and total open area of window and doors (m^2^). Each line represents the median risk over 1,000 simulations. Categories represent rooms with *x* m^2^ of total open area and quartiles above 0.

## Notes

### Competing Interest Statement

The authors have declared no competing interest.

